# The diagnostic evaluation of Convolutional Neural Network (CNN) for the assessment of chest X-ray of patients infected with COVID-19

**DOI:** 10.1101/2020.03.26.20044610

**Authors:** Syed Usama Khalid Bukhari, Syed Safwan Khalid Bukhari, Asmara Syed, Syed Sajid Hussain Shah

## Abstract

**Introduction:** The main target of COVID-19 is the lungs where it may cause pneumonia in severely ill patients. Chest X-ray is an important diagnostic test to assess the lung for the damaging effects of COVID-19. Many other microbial pathogens can also cause damage to lungs leading to pneumonia but there are certain radiological features which can favor the diagnosis of pneumonia caused by COVID-19. With the rising number of cases of COVID-19, it would be imperative to develop computer programs which may assist the health professionals in the prevailing scenario.

**Materials & Methods:** A total of two hundred and seventy eight (278) images of chest X-rays have been assessed by applying ResNet-50 convolutional neural network architectures in the present study. The digital images were acquired from the public repositories provided by University of Montreal and National Institutes of Health. These digital images of Chest X-rays were divided into three groups labeled as normal, pneumonia and COVID-19. The third group contains digital images of chest X-rays of patients diagnosed with COVID-19 infection while the second group contains images of lung with pneumonia caused by other pathogens.

**Results:** The radiological images included in the data set are 89 images of lungs with COVID-19 infection, 93 images of lungs without any radiological abnormality and 96 images of patient with pneumonia caused by other pathogens. In this data set, 80% of the images were employed for training, and 20% for testing. A pre-trained (on ImageNet data set) ResNet-50 architecture was used to diagnose the cases of COVID-19 infections on lung X-ray images. The analysis of the data revealed that computer vision based program achieved diagnostic accuracy of 98.18 %, and F1-score of 98.19.

**Conclusion:** The performance of convolutional neural network regarding the differentiation of pulmonary changes caused by COVID-19 from the other type of pneumonias on digital images of the chest X-rays is excellent and it may be an extremely useful adjunct tool for the health professionals.

## 1. Introduction

The outbreak of novel corona virus in the Wuhan city and its spread to other countries of the world has created a lot of stress and anxiety among the residents and health professionals around the globe. Corona virus is an enveloped RNA virus. There are four most common types of human corona viruses (229E, NL63, OC43, and HKU1) which causes upper respiratory tract infections such as common cold [1]. Some new strains of corona viruses like SARS-CoV, MERS-CoV, and a recently identified COVID-2019 virus evolved from corona viruses which are zoonotic in origin and these types can cause cataclysmic ailment in human being. The Middle East respiratory syndrome corona virus (MERS-CoV) is transferred to human beings from the camels. The civet cats are the source of severe acute respiratory syndrome coronavirus (SARS-CoV) and from these infected cats; it is transmitted to human beings. An outbreak of SARS-CoV has been reported from china in 2002-2003 and MERS-CoV has been identified in the patients with pneumonia in Middle East [2–4]. And now the recently identified novel coronavirus (COVID-2019) has been detected in the patients in the Wuhan city of China [5]. The mode of transmission is person to person via respiratory droplets and median incubation period is four days [6]. The disease in mild in majority of patients but in some cases severe to critical illness may occur which is characterized by breathing difficulty to respiratory failure. The severity of disease is more associated with old age. Pneumonia is the most common problem in case of severe disease. The patients with pneumonia present with the clinical features of fever, fatigue, dry cough, anorexia, myalgia, dyspnea and sputum production [7]. Radiological imaging is an important tool for the evaluation of lung diseases. In COVID-19 infection, CT scan revealed ground glass opacification which are usually bilateral and involve peripheral area of lower lobe of lungs and the radiological findings may also be present even in subclinical cases [8].

Microbial infections are the most important cause of pneumonia which include bacteria, viruses and fungi. The differentiation of pneumonia caused by viruses and particularly COVID-19 on radiological imaging is a quite difficult task which requires vast experience and strong competency in the field of radiology. The development of computer vision based programs for the detection of radiological features associated with COVID-19 on the radiological images of the lungs would be a very useful tool to facilitate heath professionals in fighting against this lethal pathogen which has affected the whole world.

The aim and objective of the present study is to evaluate the usefulness and diagnostic accuracy of computer vision-based system for the identification of radiological changes in the lungs of patients infected with COVID –19.

## 2. Materials & Methods

In the present study, two hundred and seventy eight (278) images of chest X-rays have been evaluated which were acquired from the public repositories provided by University of Montreal and National Institutes of Health [9, 10]. The digital images of Chest X-rays were divided into three groups. Group 1 included ninety three (93) digital images of chest X-ray which have no radiological abnormality and this group was labeled as Normal. Group 2 comprised of ninety six (96) digital images of chest X-rays with the radiological features of pneumonia and these patients developed pneumonia due to causes other than COVID-19 infection and this group was labeled as Pneumonia. The third group contained eighty nine (89) digital images of chest X-rays of patients diagnosed with COVID-19 infection and it was labeled as COVID-19.

For the assessment of these digital images of chest X-rays of all three groups, ResNet-50 convolutional neural network architectures has been used [11]. As our dataset was limited, we used transfer learning approach and pre-trained our models using the ImageNet data [12]. The available X-ray dataset was divided into two classes that included the Training set (80% of total dataset, i.e., 223 images), and the Testing set (20% of total dataset, i.e., 55 images). The training set is further divided into 192 images for training and 31 images for validation. We apply a number of augmentation techniques as well as a 50% dropout [13] to avoid over-fitting. A list of the the applied augmentations and their descriptions are given in Table 1. All images were resized to 224 × 224 pixels before being fed to the network.

**Table 1:** Parameters for Image augmentations

| Type of Augmentation | Description |
| --- | --- |
| horizontal_flip | A random horizontal flip is applied with probability 0.5. |
| random rotate | Inputs are randomly rotated with an angle in the range -10 to 10 degrees with 75% probability.. |
| random zoom | A random zoom in the range of 0 10 10% with 75% probability is applied. |
| random lighting | A random lightning and contrast change controlled is applied with 75% probability. |
| random warp | A random symmetric warp of magnitude between -0.2 and 0.2 is applied with 75% probability. |

We only trained the last 6-layer of the pre-trained Resnet-50 network and the remaining layers were kept fixed. We used a cyclical learning rate, ranging from 1e-6 to 1e-3 and saved the best model in every cycle using train and validation accuracy. FastAI API [14] was used to run all the algorithms on a PC with Intel-i5 with RTX-2070 GPU and 16 GB of ram.

## 3. Results

The classification results in the form of a confusion matrix for both training and test data are shown in Figure 1. Note that, only one image belonging to COVID-19 class has been classified as normal in both training and test data. Similarly, during training only one COVID-19 image was confused as non-COVID-pneumonia. This shows that the COVID-19 X-rays contain significant discriminatory information that can be extracted by a convolutional neural network. Using this extracted information, we can easily differentiate between normal, non-COVID-pneumonia and COVID-19 cases.

**Figure 1:**
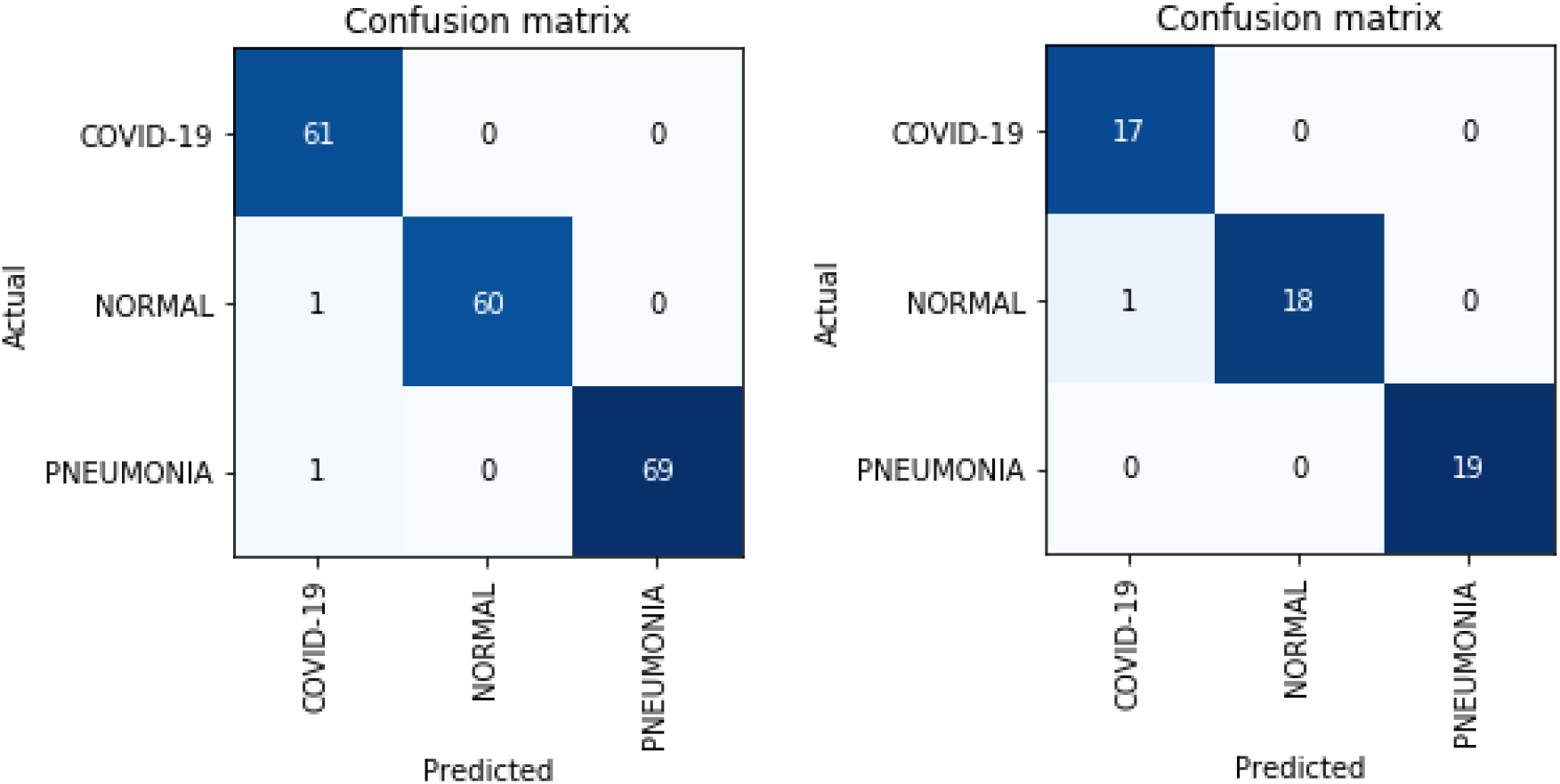
**Left**: Confusion matrix of training data. **Right**: Confusion matrix of test data.

The results in the confusion matrix are succinctly described in terms of average precision, average recall and F1-scores in Table 2 and 3. The high value of F1-score for the test data indicates the efficacy of our proposed method.

**Table 2:** Training Data Results

| Architecture | Average Precision | Average Recall | F1-Score |
| --- | --- | --- | --- |
| ResNet 50 | 0.9894 | 0.9897 | 0.9895 |

**Table 3:** Test Data Results

| Architecture | Average Precision | Average Recall | F1-Score |
| --- | --- | --- | --- |
| ResNet 50 | 0.9814 | 0.9824 | 0.9819 |

## 4. Discussion

COVID-19 has infected more than four hundred thousands persons from more than one hundred and seventy countries until now and unfortunately this number may rise in the coming days. The WHO has declared COVID-19 infection as pandemic. Extensive efforts are being done to prevent the infection and to control the damage caused by this pathogen but with the passage of time more and more cases are being diagnosed with COVID-19 infection. The most important organ attacked by COVID-19 is lung, which leads to inflammatory changes in lungs and it may cause respiratory insufficiency. The reduced oxygen supply (hypoxia) to the human cells will have deleterious effects on the human living cells and ultimately multi-organ failure may also develop in some cases which has high mortality rate. The radiological assessment of lung is important part of patient care in the severely ill patient infected with COVID-19. The evaluation of radiological imaging is a specialized job and requires the opinion of the consultant radiologist. In the prevailing current scenario, it would be imperative to develop adjunct tool which could assist the health professionals who are heavily burdened in the current situation. In this regard, one of the important areas is the interpretation of radiological images with the help of artificial intelligence. The application of deep learning machine approached may yield promising results in the detection of morphological changes in the lung of patients infected with COVID-19 on digital images of chest X-rays. The computer vision based programs may identify the subtle changes on the digital images which may not be detectable or visible to human eye. The convolutional neural network has been applied for the diagnosis of tuberculosis on the Chest X-rays and it has revealed very good results [15]. Similarly another study revealed that deep learning algorithms can help in the identification of lung opacities, enlargement of heart and pleural effusion on chest X-rays [16]. Due to the better automated feature extraction capability, the convolutional neural networks which have been trained on natural images turned out to be very successful in the classification of images [17]. The performance of convolutional neural network related to the imaging modalities with less human intervention has been found to be better than other conventional methods of machine learning [18].

In the present study, ResNet-50 convolutional neural network architectures have been applied which yielded the accuracy of 98.18 %, and F1-score of 98.19. The result of the present study are very encouraging and in near future it could be a very useful adjunct tool for the assessment of chest X-rays in case of suspected patients of COVID-19. The number of cases is one the limitation of this study. The organization and collection of the data of patient of COVID-19 from different countries of world may be planned and subjected to research for further improvement in the diagnostic modalities.

## 5. Conclusion

The performance of convolutional neural network regarding the differentiation of pulmonary changes caused by COVID-19 from the other type of pneumonias on digital images of the chest X-rays is extremely good. It may be a very useful adjunct tool for the identification of lung changes caused by COVID-19 infection.

## Data Availability

the data is acquired from the public repositories provided by University of Montreal and National Institutes of Health

https://github.com/ieee8023/covid-chestxray-dataset

https://www.nih.gov/news-events/news-releases/nih-clinical-center-provides-one-largestpublicly-available-chest-x-ray-datasets-scientific-community

## Acknowledgements

The authors are thankful to University of Montreal and National Institutes of Health for making the data public and allow us to conduct this research. The authors are also thankful to DockCloud company for letting us use their hardware and services.

## Scientific Responsibility Statement

The authors declare that they are responsible for the article’s scientific content including study design, data collection, analysis and interpretation, writing, some of the main line, or all of the preparation and scientific review of the contents and approval of the final version of the article.

## Animal and human rights statement

All procedures performed in this study were in accordance with the ethical standards of the institutional and/or national research committee and with the1964 Helsinki declaration and its later amendments or comparable ethical standards. No animal or human experimental studies were carried out by the authors for this article.

## Funding

Nil

## Conflict of interest

None of the authors received any type of financial support that could be considered potential conflict of interest regarding the manuscript or its submission.

## Notes

### Competing Interest Statement

The authors have declared no competing interest.

### Funding Statement

no funding was received

## References

[1] P. Habibzadeh, E. K. Stoneman, The Novel Coronavirus: A Bird’s Eye View, The International Journal of Occupational and Environmental Medicine 11 (2) (2020) 65–71. doi:10.15171/ijoem.2020.1921. URL https://dx.doi.org/10.15171/ijoem.2020.1921

[2] J. Cui, F. Li, Z.-L. Shi, Origin and evolution of pathogenic coronaviruses, Nature Reviews Microbiology 17 (3) (2019) 181–192. doi:10.1038/s41579-018-0118-9. URL https://dx.doi.org/10.1038/s41579-018-0118-9

[3] N. S. Zhong, B. J. Zheng, Y. M. Li, Epidemiology and cause of severe acute respiratory syndrome (SARS) in Guangdong, People’s Republic of China, The Lancet 362 (2003) 1353–1358.

[4] A. M. Zaki, S. van Boheemen, T. M. Bestebroer, A. D. Osterhaus, R. A. Fouchier, Isolation of a Novel Coronavirus from a Man with Pneumonia in Saudi Arabia, New England Journal of Medicine 367 (19) (2012) 1814–1820. doi:10.1056/nejmoa1211721. URL https://dx.doi.org/10.1056/nejmoa1211721

[5] N. Zhu, D. Zhang, W. Wang, A novel coronavirus from patients with pneumonia in China, N Engl J Med (2019).

[6] W. J. Guan, Z. Y. Ni, Y. Hu, W. H. Liang, C. Q. Ou, J. X. He, Clinical Characteristics of Coronavirus Disease 2019 in China, N Engl J Med (2020).

[7] D. Wang, B. Hu, C. Hu, F. Zhu, X. Liu, J. Zhang, Clinical Characteristics of 138 Hospitalized Patients With 2019 Novel Coronavirus-Infected Pneumonia in Wuhan (2020).

[8] H. Shi, X. Han, N. Jiang, Y. Cao, O. Alwalid, J. Gu, Y. Fan, C. Zheng, Radiological findings from 81 patients with COVID-19 pneumonia in Wuhan, China: a descriptive study, The Lancet Infectious Diseases (20) (2020) 30086–30090. doi:10.1016/s1473-3099(20)30086-4. URL https://dx.doi.org/10.1016/s1473-3099(20)30086-4

[9] J. P. Cohen, Covid-19 image data collection, https://github.com/ieee8023/covid-chestxray-dataset (2020).

[10] N. I. of Health Clinical Center, Nih chest x-ray dataset of 14 common thorax disease categories. URL https://www.nih.gov/news-events/news-releases/nih-clinical-center-provides-one-largest-publicly-available-chest-x-ray-datasets-scientific-community

[11] K. He, X. Zhang, S. Ren, J. Sun, Deep residual learning for image recognition, in: Proceedings of the IEEE conference on computer vision and pattern recognition, 2016, pp. 770–778.

[12] O. Russakovsky, J. Deng, H. Su, J. Krause, S. Satheesh, S. Ma, Z. Huang, A. Karpathy, A. Khosla, M. Bernstein, Imagenet large scale visual recognition challenge, International journal of computer vision 115 (3) (2015) 211–252.

[13] N. Srivastava, G. Hinton, A. Krizhevsky, I. Sutskever, R. Salakhutdinov, Dropout: a simple way to prevent neural networks from overfitting, The journal of machine learning research 15 (1) (2014) 1929–1958.

[14] J. Howard, et al., fastai, https://github.com/fastai/fastai (2018).

[15] F. Pasa, V. Golkov, F. Pfeiffer, D. Cremers, D. Pfeiffer, Efficient Deep Network Architectures for Fast Chest X-Ray Tuberculosis Screening and Visualization, Scientific Reports 9 (1) (2019). doi:10.1038/s41598-019-42557-4. URL https://dx.doi.org/10.1038/s41598-019-42557-4

[16] R. Singh, M. K. Kalra, C. Nitiwarangkul, J. A. Patti, F. Homayounieh, A. Padole, P. Rao, P. Putha, V. V. Muse, A. Sharma, Deep learning in chest radiography: detection of findings and presence of change, PloS one 13 (10) (2018).

[17] S. Rajaraman, I. Kim, S. K. Antani, Detection and visualization of abnormality in chest radiographs using modality-specific convolutional neural network ensembles, PeerJ 8 (2020) e8693–e8693. doi:10.7717/peerj.8693. URL https://dx.doi.org/10.7717/peerj.8693

[18] I. Kim, S. Rajaraman, S. Antani, Visual interpretation of convolutional neural network predictions in classifying medical image modalities, Diagnostics 9 (2) (2019) 38.

